# Steep HIV incidence declines among female sex workers in Cotonou, Benin: reconstructing finely stratified HIV incidence estimates from HIV prevalence 1993-2022

**DOI:** 10.64898/2026.07.21.26358335

**Authors:** Oliver Stevens, Rebecca L Anderson, Souleymane Diabate, Jeffrey W. Imai-Eaton

**Affiliations:** MRC Centre for Global Infectious Disease Analysis, School of Public Health, Imperial College London, London, United Kingdom; Département de médecine sociale et préventive, Université Laval, Canada; Center for Communicable Disease Dynamics, Department of Epidemiology, Harvard T. H. Chan School of Public Health, Boston, MA, USA

## Abstract

**Background:** HIV prevalence among female sex workers in Benin has declined steeply over the last thirty years, driven by successful HIV prevention and treatment programming among sex workers and their clients. To maintain progress in an era of constrained funding, HIV prevention programming should be targeted towards those most at risk of HIV seroconversion. We present a model-based approach that produces finely stratified HIV incidence estimates from widely available HIV prevalence data from serial cross-sectional surveys among female sex workers.

**Methods:** We analysed participant-level data from twelve cross-sectional surveys among FSW in Cotonou, Benin from 1993-2022. We created a compartmental model representing women transitioning into and out of sex work and acquiring HIV. The model was stratified by single year of age, duration at-risk, calendar year, and time since HIV seroconversion and calibrated to HIV prevalence by single-year age and duration at-risk, and age and duration distribution data.

**Results:** HIV prevalence among sex workers aged 15-49 in Cotonou declined from 44.9% (95%CI 41.2-48.9) in 1995 to 8.6% (95%CI 6.4-11.8%) in 2022 (Figure 1). HIV incidence declined by 86% (95%CI 82-90%) between 1994 and 2013 from 19.7/100py (95%CI 9.5-26.9) to 2.5/100py (95%CI 1.2-3.5), and remained stable thereafter through 2022 (Figure 2A). Throughout the study period, new infections were concentrated among women who recently started selling sex: in 2022, 72% of infections occurred in the first year of sex work (95%CI 64-81%; Figure 2B). Incidence patterns by age varied less than by duration and was similar at around 1.5/100py for all sex workers under age 30, rising to 4/100py among those over age 40. Modelled incidence exceeded empirical cohort estimates, highlighting sensitivity to assumptions about HIV prevalence at sex work initiation, episodic sex work, duration misclassification, and population size.

**Conclusion:** We estimated large declines in HIV incidence among sex workers in Benin from 1994-2013, but slower progress from 2013-2022. New sex workers across all age groups, who may be weakly linked to programmes, should be the highest priority for HIV prevention interventions and community outreach efforts. This modelling approach to empirically monitor incidence among sex workers should be applied across high burden epidemic settings with serial survey data to guide prevention efforts, including the allocation of limited provision of long-acting pre-exposure prophylaxis.

---

Estimates of HIV incidence are key to monitoring the HIV epidemic, have formed central components of strategic information portfolios since the early epidemic, and are now urgently required among key populations to target limited provision of long-acting HIV prophylaxis. Empirical incidence studies, mostly commonly prospective cohort studies, require longitudinal follow-up and accordingly are expensive and logistically challenging to implement. Implementation is further complicated among sex workers as criminalisation and frequently moving for work hamper follow-up and introduce heterogeneity into cohorts that may lead to biased estimates of incidence.^1^ Our recent systematic review identified only 32 incidence studies among female sex workers conducted in sub-Saharan Africa across the entire epidemic.^1^ Meta-analysis of these incidence studies estimated incidence among sex workers was 8 times higher than among subnational location and age matched total population women. Instead, model-based estimates calibrated to cross-sectional behavioural and HIV surveillance data are commonly used to estimate HIV incidence.^2–4^

Transmission dynamic models that seek to reconstruct the complete epidemic history among all subpopulations are necessarily data demanding.^5–8^ In the case of key populations, where data are sparse, this either restricts their application to the most data rich settings or results in assumption-heavy approaches. Model representations of sex work behaviour are often granular and may incorporate both paying and non-paying partners and their HIV prevalence; frequency of clients, vaginal, and anal sex; and all associated HIV prevention and treatment strata. The absence of direct client population size estimates from most settings is a large source of uncertainty in such models and must be inferred from FSW sexual partner data.^7^ Estimates of the proportion of men who buy sex from total population surveys vary widely and are assumed to be under-estimated.^7,9^

Instead, we present a statistical model that estimates HIV incidence among female sex workers using single-year age- and duration-stratified cross-sectional survey data. Several methods have sought to reconstruct incidence from serial cross-sectional measures of prevalence among the total population in sub-Saharan Africa where, setting aside migration, mortality is the only opportunity for individuals to leave the population. Catalytic-based approaches calculate incidence from age-specific prevalence under equilibrium conditions. As the age pattern of prevalence is assumed to be constant, prevalence reflects the cumulative exposure to incidence. Gregson et al. detail a catalytic method which fits a functional incidence form to age-specific ANC prevalence data under the assumption of steady state conditions.^10^ Several elaborations on this catalytic model have been developed for application in sub-Saharan Africa. Williams et al. specified a separable force of infection which could change over time but maintained a constant age pattern to estimate incidence in South Africa’s growing epidemic.^11^ Mossong et al. jointly estimated HIV incidence and survival since seroconversion from HIV prevalence alone applied to the same serial prevalence data in Williams et al.

Gregson et al. also detail a simple cohort-based approach comparing prevalence among neighbouring age groups at a single point in time, estimating incidence as the number of infections required to obtain the change in cohort prevalence. This produced similar incidence estimates to their catalytic model. However, both Gregson methods and the catalytic models assume constant age patterns of prevalence over time, an unrealistic assumption for much of the epidemic, nor link serial measures of prevalence as a longitudinal cohort. Instead, using serial cross-sectional measures of prevalence that track hypothetical cohorts over time allows us to estimate incidence in non-steady state conditions: comparing prevalence at time *t*in age group (*a*_0_,*a*_1_) with prevalence at time (*t*+ *x*) in age group (*a*_0_ + *x,a*_1_ + *x*). This produces an incidence estimate for each inter-survey period. Hallett et al. detail these longitudinal ‘demographic accounting’ methods, accounting for the depleting effect of HIV and non-HIV mortality among prevalent PLHIV between surveys.^2,3^ This method may be applied in any epidemic state and yields comparable incidence estimates to empirical observations from population-based cohort studies.

Finally, transmission dynamic models fit a continuous transmission or incidence rates within a compartmental model framework and optimise for best fit to observed prevalence data. These include a wide range of models including EPP-ASM ^4^, the incidence model used within the UNAIDS HIV estimates in sub-Saharan Africa which uses a basic epidemic structure, to finely behaviourally stratified models that have been used for key population estimation. All methods require external estimates of mortality or survival since seroconversion, which represent the least certain model inputs: routine mortality surveillance in sub-Saharan Africa is very rare, estimates of survival since seroconversion are derived from studies in the pre-treatment era, and mortality estimates among those on ART confounded by CD4 count eligibility changes and treatment interruption.^12–14^

Calculating incidence among sex workers from changes in cross-sectional prevalence over time is complicated by the additional dynamic of initiating and ceasing risk behaviour, often referred to as ‘turnover’.^15^ If women sell sex for, on average, five years, 20% of the population exit sex work each year. Younger women with lower HIV prevalence initiate sex work and experience high risk of HIV seroconversion during their sex work career. Older women, now with higher HIV prevalence, cease selling sex and re-enter the total population. As the population outflow has higher prevalence than the inflow, HIV incidence can remain high over time despite HIV prevalence remaining constant or declining. In settings where HIV prevalence among sex workers is declining over time, estimating HIV incidence without accounting for population turnover leads to implausibly low, or even negative, estimates.^16^ Including duration at-risk (*d*) within demographic accounting methods acts as an additional dimension by which we may estimate incidence among sex workers: we can compare prevalence at *a*_0_ and (*a*_0_ + *x*) as in Hallett et al., and additionally and jointly compare prevalence at *d*_0_ and (*d*_0_ + *x*).

This analysis seeks to create a statistical model that directly estimates annual HIV incidence among sex workers from data on HIV prevalence by sex worker cohort in successive surveys, without relying on assumptions about transmission dynamics or sexual mixing.

## 1.2 Data and methods

Participant-level data were available from twelve cross-sectional surveys among female sex workers conducted in Cotonou, Benin from 1993-2023 that measured age, HIV prevalence, and reported duration in sex work, among other information. A compartmental model was created representing women transitioning in and out of sex work and acquiring HIV infection. Excess HIV mortality was calculated using a hybrid approach of survival since seroconversion and age-specific mortality rates.

HIV incidence stratified by single-year age, sex work duration and was modelled using mixed-effect Bayesian regression. The average duration of sex work was estimated and allowed to vary over time. The model was calibrated to HIV prevalence by age and duration, and the count distribution of women selling sex by age and duration.

### 1.2.1 Data

The Benin National AIDS Council, in collaboration with a Canadian development and research partnership, has supported *Projet Sida-2* and *Dispensaire IST*, a clinic in Cotonou since the early 1990s. *Dispensaire IST* provides HIV services, including prevention, testing, and treatment, to female sex workers. The programme routinely implemented cross-sectional surveys to monitor the epidemic among sex workers (Table 6.1). Surveys initially recruited using convenience samples of programme attendees, in conjunction with outreach to known sex work hotspots to encourage programme attendance (1993, 1995, 1998, and 2008). Later surveys used hotspot-based sampling (2002, 2005, 2011, 2015, 2017, and 2022), and from 2011 sampling was expanded to cities throughout Benin. This analysis is restricted to data collected in Cotonou.

**Table 6_1:**
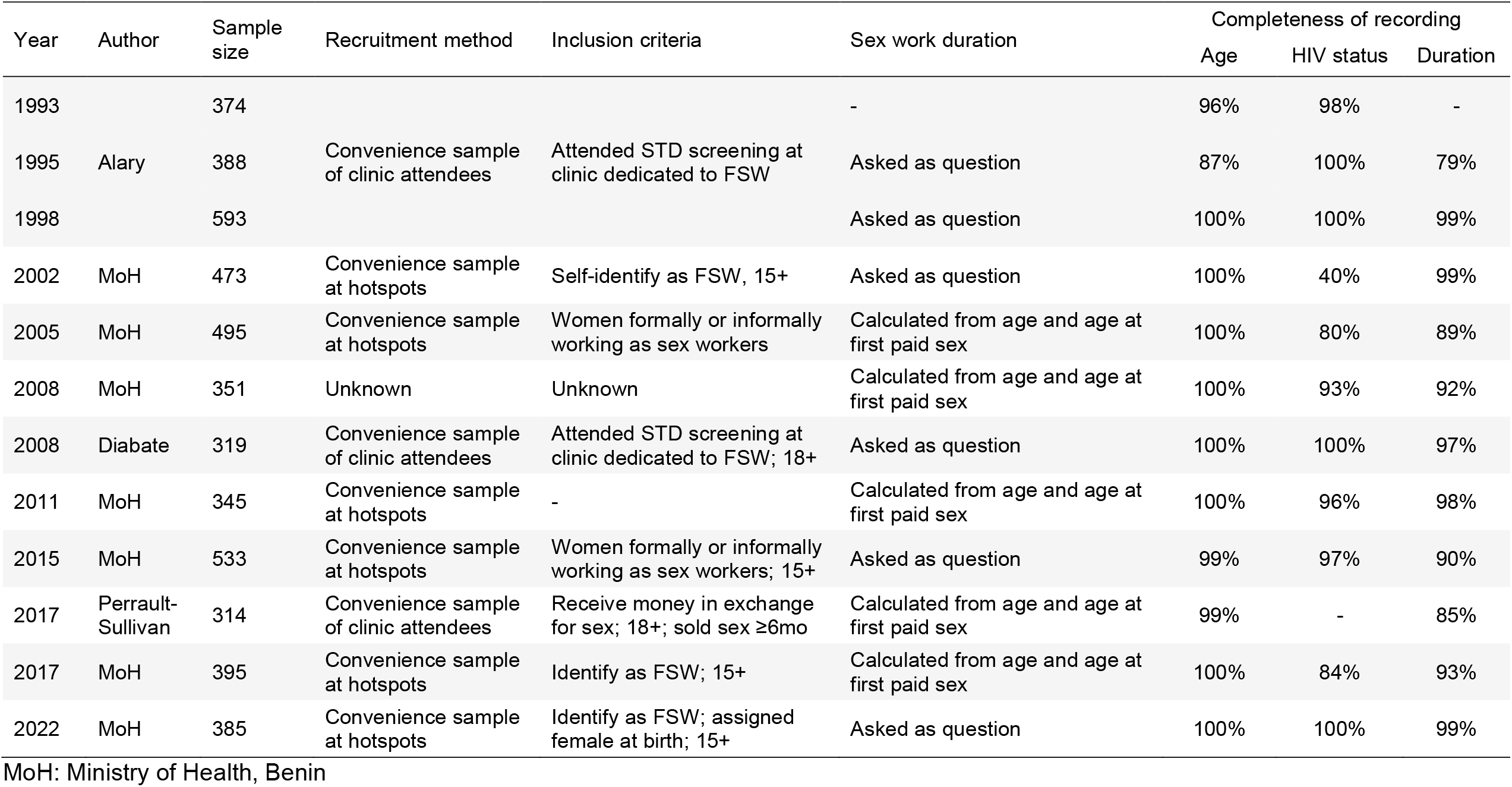
Surveillance studies among female sex workers in Cotonou, Benin.

### 1.2.2 Compartmental model structure

HIV-negative female sex workers were stratified by calendar year {*t* ∈ 1993: 2022},single year of age {*a* ∈ 15,16 . . 59,60+}, and single year of duration selling sex {*d* ∈ <1,1,2 . . 19,20+} (Figure 6.2). HIV-positive sex workers were further stratified by single year since seroconversion {*s* ∈ <1,1,2 . . 14,15+}. The minimum age that sex workers could begin selling sex was age 15, therefore 15 year olds must all be in the first year of selling sex, 16 year olds in the first two years etc.

#### 1.2.2.1 Demographic progression, turnover, and mortality

Each year, a proportion of female sex workers cease to sell sex, 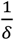 where *δ* is the mean duration of sex work. The log mean duration of selling sex over time was modelled with an intercept representing the average duration (*β*_*δ*0_) and smooth temporal effect (*γ*_*δt*_). The temporal component was modelled as a cubic B-spline with *K* = 7 knots equally spaced every five calendar years from *t* ∈ {1993,1998 … 2022}. The spline coefficients were penalised with a first-order autoregression model, with the autocorrelation parameter fixed at *ϕ* = 0.95 to enforce smoothness.

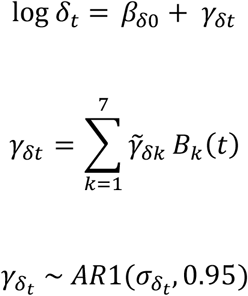

A further proportion die each year (*μ*). Non-HIV mortality among sex workers was assumed to be the same as that among age-matched women, and was extracted from the Benin Spectrum file by five-year age group and year. It was interpolated to single year of age using Beers ordinary interpolation 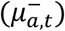. All-cause mortality rates among people living with HIV stratified by age and year since HIV seroconversion in the pre-treatment era were estimated by Todd et al.^17^ (Figure 6.3A). Non-HIV mortality was subtracted from the Todd rates to estimate excess AIDS mortality by infection duration in the pre-treatment era 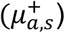. To estimate AIDS mortality in the treatment era without an estimate of the number of sex workers on ART, an ART mortality rate ratio by age and year (*MRR*_*a,t*_) since treatment became available (*ART*_*t*_) was calculated by dividing total population AIDS mortality rates relative to rates in the year that treatment became available (*ART*_*t*_= 0) for all countries in sub-Saharan Africa (Figure 6.3B). Excess AIDS mortality rates stratified by age, time since seroconversion, and year were calculated by multiplying 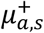 and the mortality rate ratio (Figure 6.3C):

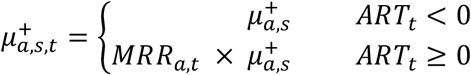

Surviving sex workers age by one year, increase one year of duration selling sex, and HIV-positive sex workers increase one year since HIV seroconversion, such that:

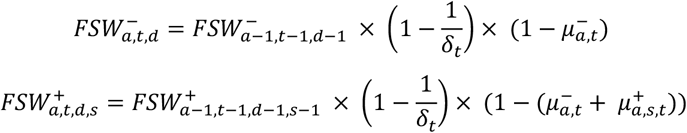

The final age, duration, and seroconversion indices are cumulative and represent those aged ≥ 60, selling sex for ≥20 years, or having seroconverted ≥15 years ago.

#### 1.2.2.2 Recruitment of new sex workers

Total population women initiate sex work at rate *ψ* given by an intercept and a smooth AR1-penalised B-spline pattern over time:

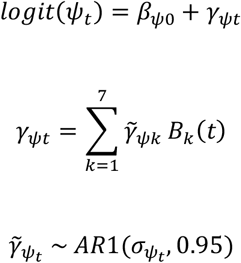

The annual sex worker population size is calculated fixed at 1% of the total 15-49 female population), and the appropriate number of sex workers is recruited into duration group *d* = 0 to reach this population size (*X*). Demographic progression or HIV dynamics among the total population were not modelled. Instead, the total number of women by year, age, and HIV status were extracted from the Spectrum file for Benin from which new sex workers were recruited. The proportion *π*_*a*_ entering sex work at each age *a* was modelled as a gamma distribution and is estimated from the data and allowed to change over time:

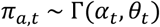

The gamma distribution was paramaterised in terms of a mean (*μ*) and a standard deviation (*σ*):

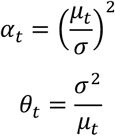

Where the log mean age of entering sex work was specified with an intercept and a smooth AR1-penalised B-spline temporal effect:

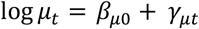

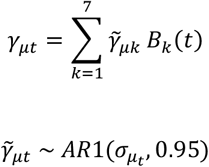

*X*_*a*.*t*_(the number of women initiating sex work) is given by the initiation rate (*ψ*) multiplied by the total female population size and the age distribution of new sex workers (*π*):

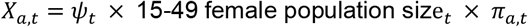

The annual initiation rate is identified by summing HIV-negative, HIV-positive, and newly recruited FSW and applying a soft constraint against the target population size, fixed at 1% of the total female population:

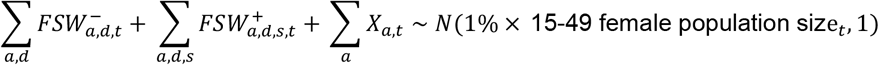

To reflect that women initiating sex work were likely at higher risk of HIV acquisition before formal entry into sex work^18^, the prevalence of women entering sex work is adjusted from the total population prevalence by age and time. The age-specific odds of being HIV-positive was multiplied by an odds ratio that varies over time which is estimated from the data (*ι*_*t*_). This odds ratio was constrained to be between 1 and 5.

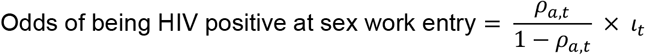

No data exist in Benin on the time since seroconversion for women who are HIV-positive at sex work entry. A recent phylogenetic study in Kenya estimated that prevalent sex workers seroconverted on average 1.5 years before formal entry into sex work. ^18^ This model assumes a fixed distribution of 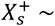 Weibull(2,1.4) across age and time, which assumes prevalent positives have recently seroconverted (80% of the mass <2 years).

#### 1.2.2.3 Modelling HIV incidence

HIV incidence (*λ*) was then applied to the HIV-negative FSW population, and new infections added to the HIV-positive population at *s* = 0.

Incidence was assumed to vary smoothly over time, age, and duration. Additional flexibility permitted incidence to vary differentially and smoothly by age over time. The logit annual probability of acquiring HIV infection by time, age, and duration was modelled with an intercept (*β*_*λ*0_) and AR1-penalised B-splines with *K* knots over age (*γ*_*λa*_; *K* = 12), time (*γ*_*λt*_; *K* = 9), duration of selling sex (*γ*_*λd*_; *K* = 7), and an interaction between age and time (*η*_*λa,t*_).

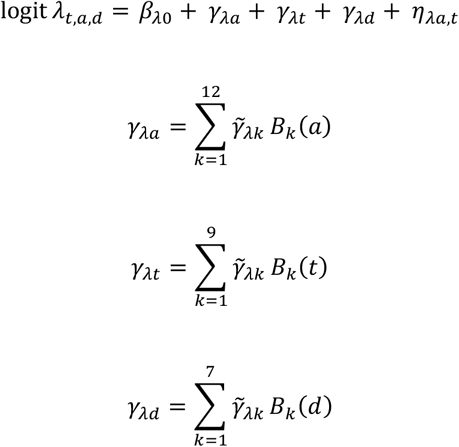

Incidence rate by duration of sex work was enforced to decline monotonically:

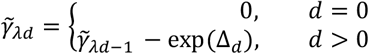

where the log-differences between duration knots (Δ_*d*_) were penalised with an AR1 process.

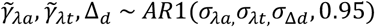

The age-time interaction was specified as a separable interaction of the smooth main age and time effects to permit differential age patterns of HIV incidence over time:

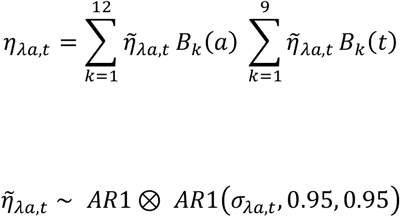

#### 1.2.2.4 Initialising the model

The model did not estimate the entire epidemic; the female sex worker population was initialised in 1993 by age, duration and HIV prevalence. The initial age distribution of sex workers was given by a gamma distribution with shape and scale parameters estimated from the data: *a*_*init*_∼ Γ(*α,θ*). Sex workers at each age were distributed by duration selling sex such that:

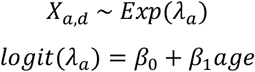

Finally, sex workers by age and duration were distributed by HIV status. To achieve HIV prevalence patterns consistent with HIV incidence trajectories, initial prevalence in 1993 was calculated from HIV incidence and mortality in the first year of the simulation under the assumption of steady-state conditions. The prevalence at age (*a* + 1) and duration (*d* + 1) is the prevalence at age *a* and duration *d* less mortality:

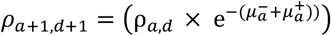

plus those who seroconvert and avoid HIV-negative mortality:

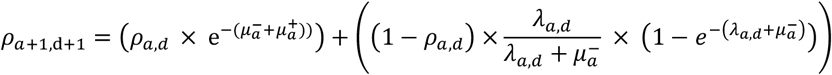

### 1.2.3 Priors and constraints

Fixed effects had weakly informative priors: *β* ∼ *N*(0,31.6)

Standard deviations of random effects had half normal priors: *σ* ∼ *N*(0,2.5)

All smoothing random effects were given soft sum-to-zero constraints to aid identifiability.

### 1.2.4 Likelihood for FSW survey prevalence and duration data

#### 1. Prevalence

The number of HIV-positive FSW by age and duration was calibrated survey-observed (*x*) prevalence (*ρ*_*a,d,x,t*_):

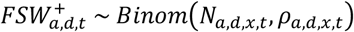

Survey respondents who did not report a duration of sex work were incorporated into calibration by aggregating over all duration categories, assuming that the missing durations were missing at random:

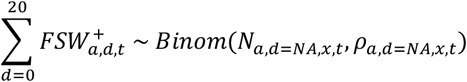

#### 2. Age-duration distribution

The modelled proportion of FSW within each age-duration category was calculated in each year:

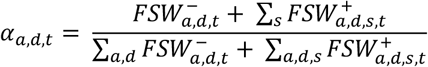

An observation model adjusted for under-recruitment of FSW at short durations of selling sex (<1 and 1-2 years), relating the true age-duration distribution to that observed in each survey 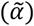. Underenumeration (*θ*) was estimated as a multiplicative factor on the natural scale. Fixed effects at *d* = 0 and *d* = 1 captured the average global under-enumeration at short durations, and survey iid effects *ω*_*x*_ permitted survey-specific deviation from each global average. Underenumeration at *d* = 0 was enforced to be more severe than at *d* = 1 such that:

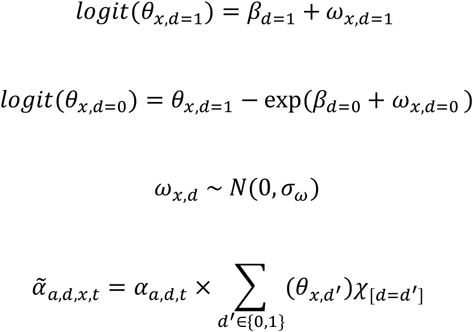

The bias-adjusted proportions were normalised, converted back to counts, and calibrated to the age-duration distribution in each survey:

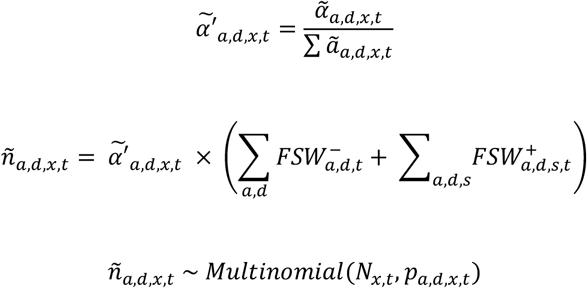

### 1.2.5 Validation

Empirical observations of HIV incidence among female sex workers in Benin were available from two studies. Van Damme et al.^19^ observed 10 new infections from 1998-2002 (8.3/100py 95%CI 3.4, 13.2) and Diabaté et al.^20^ observed 6 new infections from 2008-2012 (1.4/100py 95%CI 0.3, 2.5). Both studies recruited women aged 18 or over. Incidence estimates for ages 18+ in 2000 and 2010, the study midpoints, were compared to empirical observations.

### 1.2.6 Sensitivity analysis

Several sensitivity analyses were conducted:

1. Removing the under-enumeration observation model when calibrating to the age-duration distribution;
2. Increasing the population size estimate proportion to 2% in 2020, and 1.5% in 2021, returning to 1% in 2022 to simulate a COVID-19 pandemic shock growth in sex work. Fixed effects were added into the incidence and initiation models, estimated only in years 2020 and 2021:

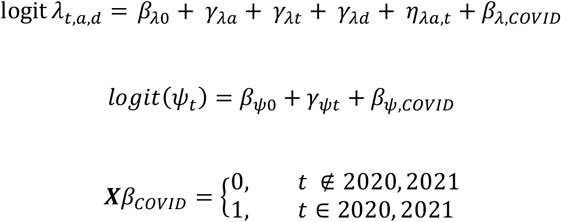
3. Alternative specifications for the odds ratio that increases HIV prevalence at sex work initiation (*ι*_*t*_): constant over time at *ι*_*t*_ ∈ {1,3,5,10} and varying over time with no upper bound (main results constrain *ι*_*t*_between 1 and 5);
4. An additional fixed effect in the incidence model to capture very high incidence at *d* = 0:

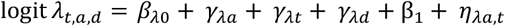

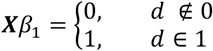

### 1.2.7 Model implementation

Data preparation was conducted in R and the model was implemented in C++ via the R package Template Model Builder (TMB). One thousand samples were drawn from the joint posterior distribution, from which the posterior mean, standard deviation, and quantile-based 95% CI were calculated for each output indicator.

## 1.3 Results

### Descriptive data statistics

4,966 female sex workers participated in 12 surveys conducted between 1993-2022 that measured serological HIV status (N surveys = 11; median completeness 96.6%, interquartile range [IQR] 88.5-99.5%), age (N=12; 99.5% IQR 99.3-100%), and duration of selling sex (N = 11; 92.7% IQR 89.7-98.4; Table 6.1). Six surveys included a question on duration of sex work, while duration was calculated for five surveys using current age and reported age at first paid sex. All surveys used convenience-based sampling, of which five used clinic-based recruitment. HIV status data from the 1993 survey was excluded from model calibration due to implausible trends in HIV prevalence by age.

### Modelled HIV prevalence estimates

HIV prevalence among all sex workers declined consistently from its peak in 1995 at 44.9% (95CrI 41.2-48.9) to 8.6% (95CrI 6.4-11.8%) in 2022 (Figure 6.1A). Prevalence has declined among all age groups, with largest relative declines among the younger age groups (approximately eight-fold decline in 15-19 and 20-24 year olds; Figure 6.1B). In 2022, HIV prevalence remains strongly correlated with age, increasing by *circa* 50% in each successive five-year age group between ages 15 and 49. Trends in HIV prevalence by sex work duration were less consistent (Figure 6.1C). HIV burden increased in the first 3-5 years of sex work and then plateaued, though estimates at long durations of sex work were weakly informed by data, particularly in the pre-treatment era.

**Figure 6_1:**
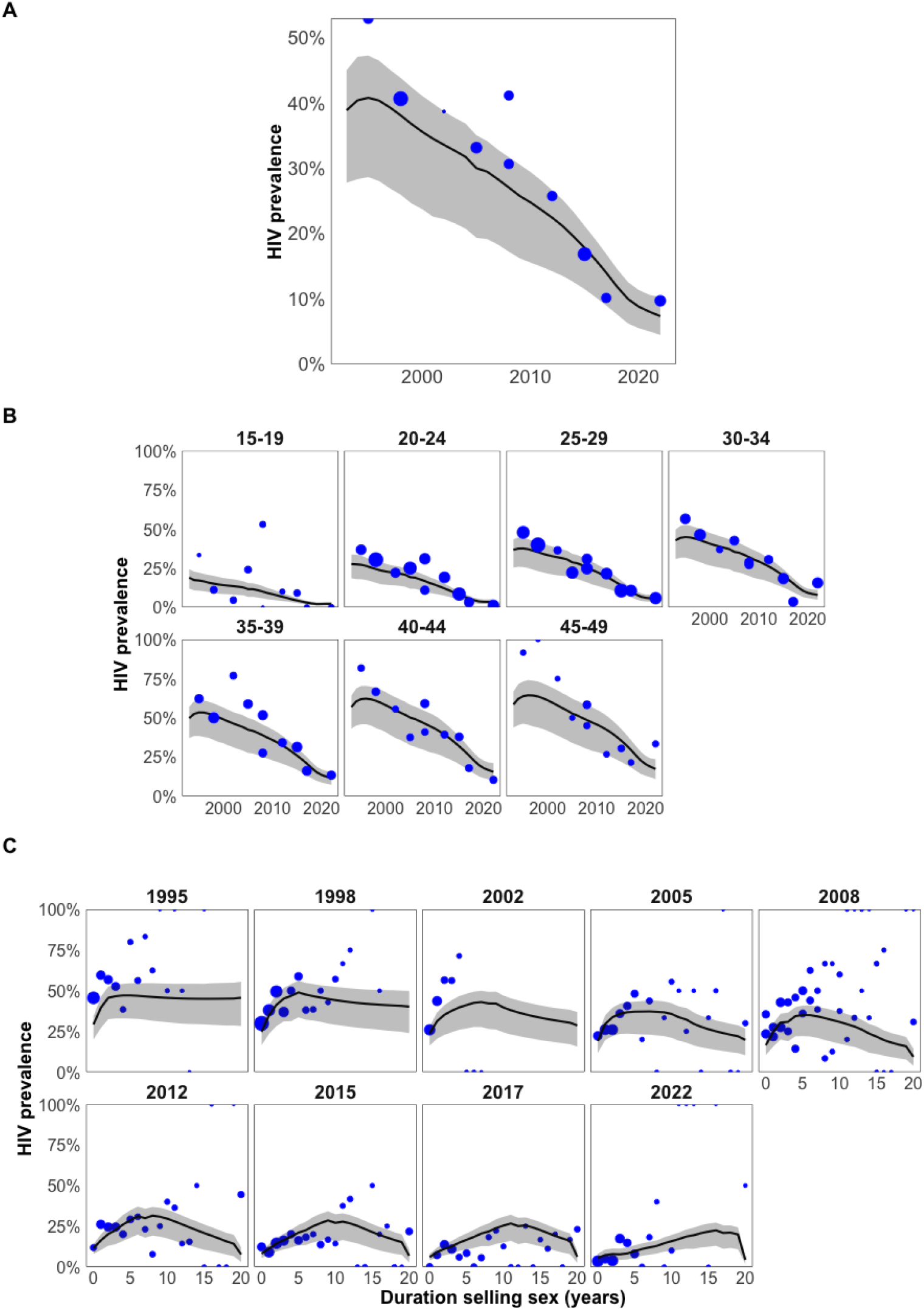
HIV prevalence. (A) All sex workers over time; (B) By five-year age group over time; (C) All sex workers by duration of selling sex, shown for years of survey. Model estimates represented by black line with 95% uncertainty range in grey shading. Survey data shown in blue points, sized by sample size.

**Figure 6_2:**
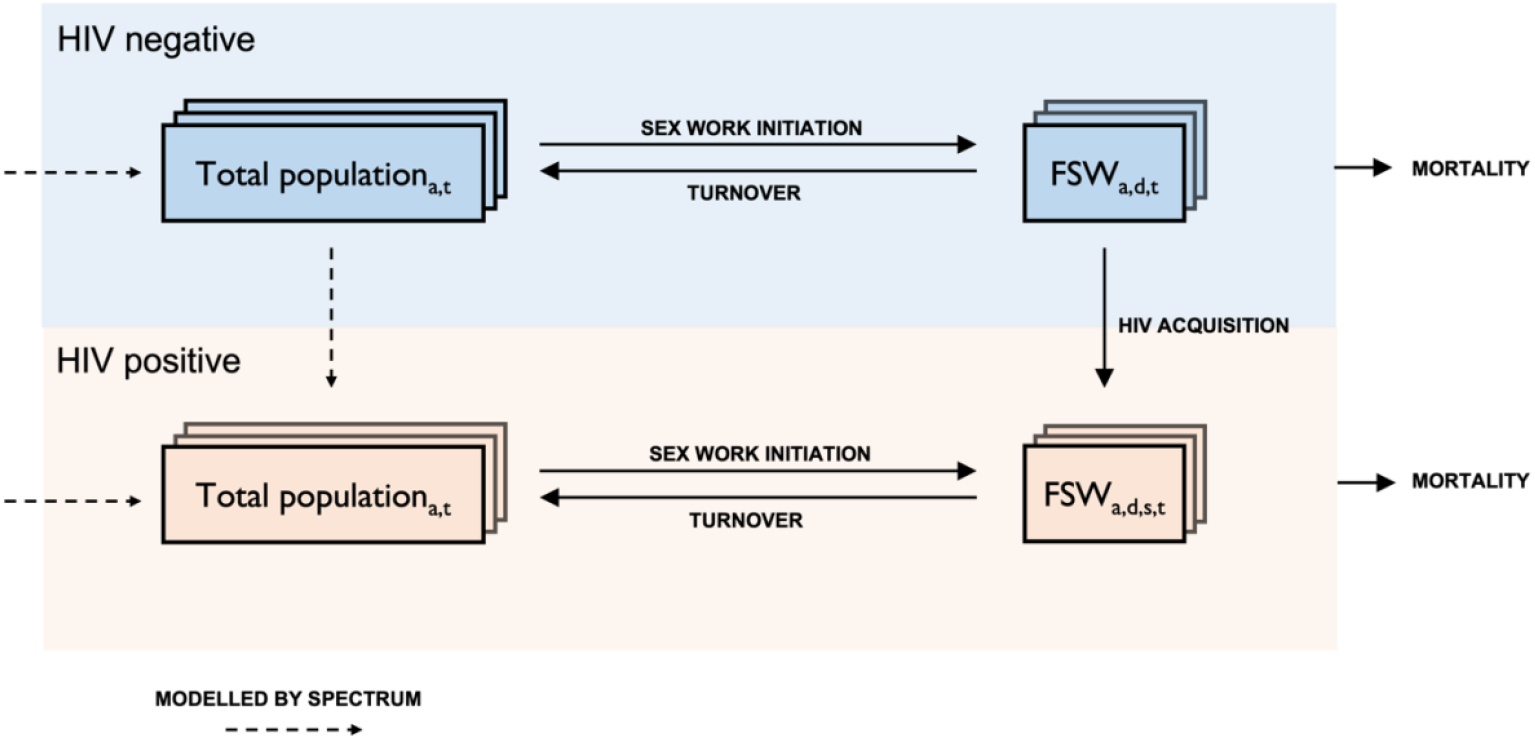
Compartmental model schematic diagram.

### Modelled HIV incidence estimates

HIV incidence among sex workers aged 15-49 has declined from 19.7/100py (95CrI 9.5-26.9) in 1994 to 2.5/100py (1.2-3.5) in 2013 where it has remained relatively stable 2014-2022 (Figure 6.2A). HIV incidence was concentrated at the lowest durations, with half of cumulative incidence and 81% of new infections occurring within the first two years of selling sex (Figure 6.2B and C). Incidence among 15-29 has converged over time to around 1.5/100py in 2022 (Figure 6.2D), but remains higher in older age groups (Figure 6.2E). In 2022, 60% of new infections occurred among those aged 20-34 (Figure 6.2F).

**Figure 6_2a:**
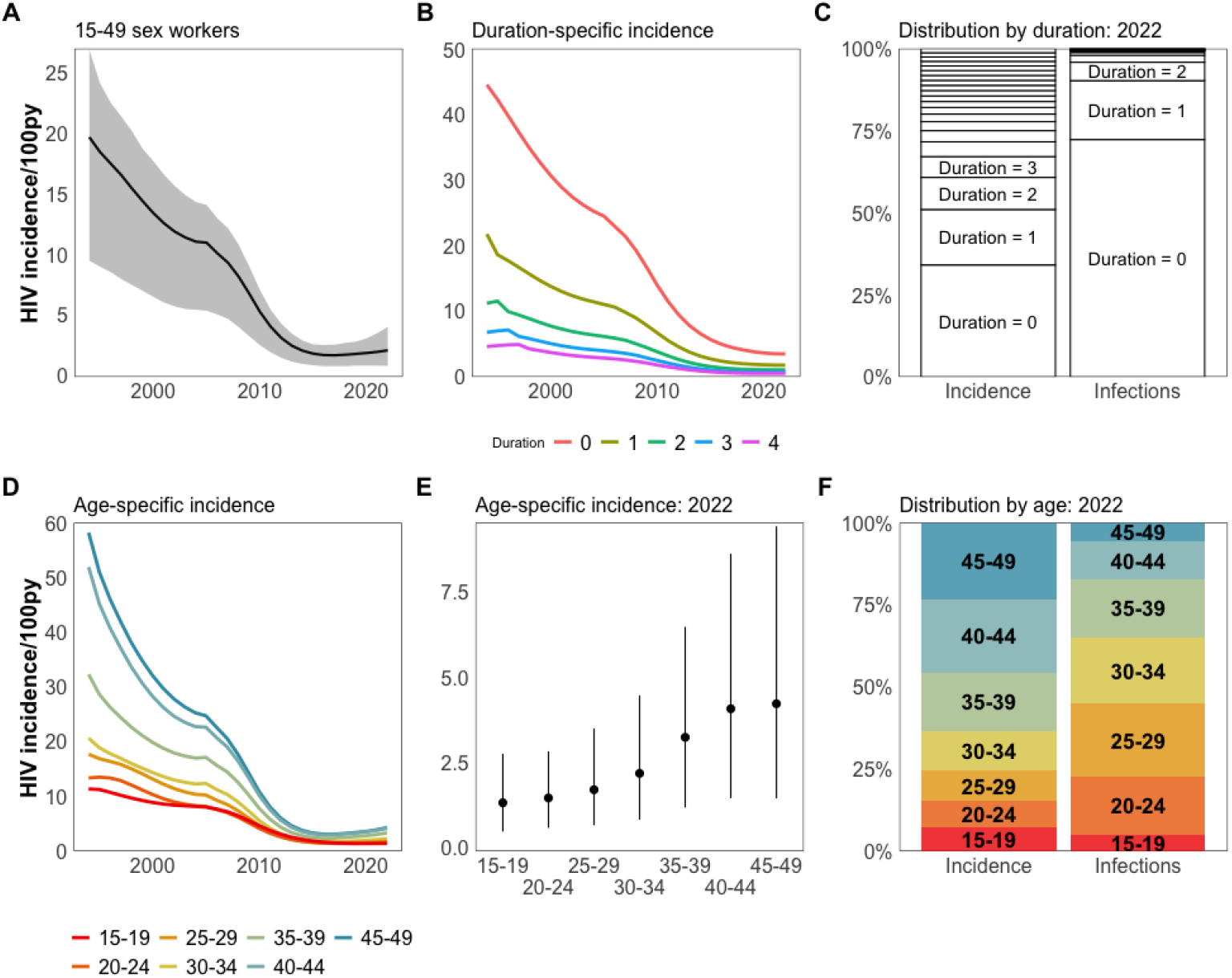
Estimates of HIV incidence. (A) Among 15-49 sex workers over time; (B) By duration for the first five years of sex work over time; (C) Distribution of incidence and new infections by duration in 2022; (D) Age-specific incidence over time; (E) Age-specific incidence in 2022; (F) Distribution of incidence and new infections by five-year age group in 2022. 95% credible interval omitted from (B) and (D) for clarity.

**Figure 6_3:**
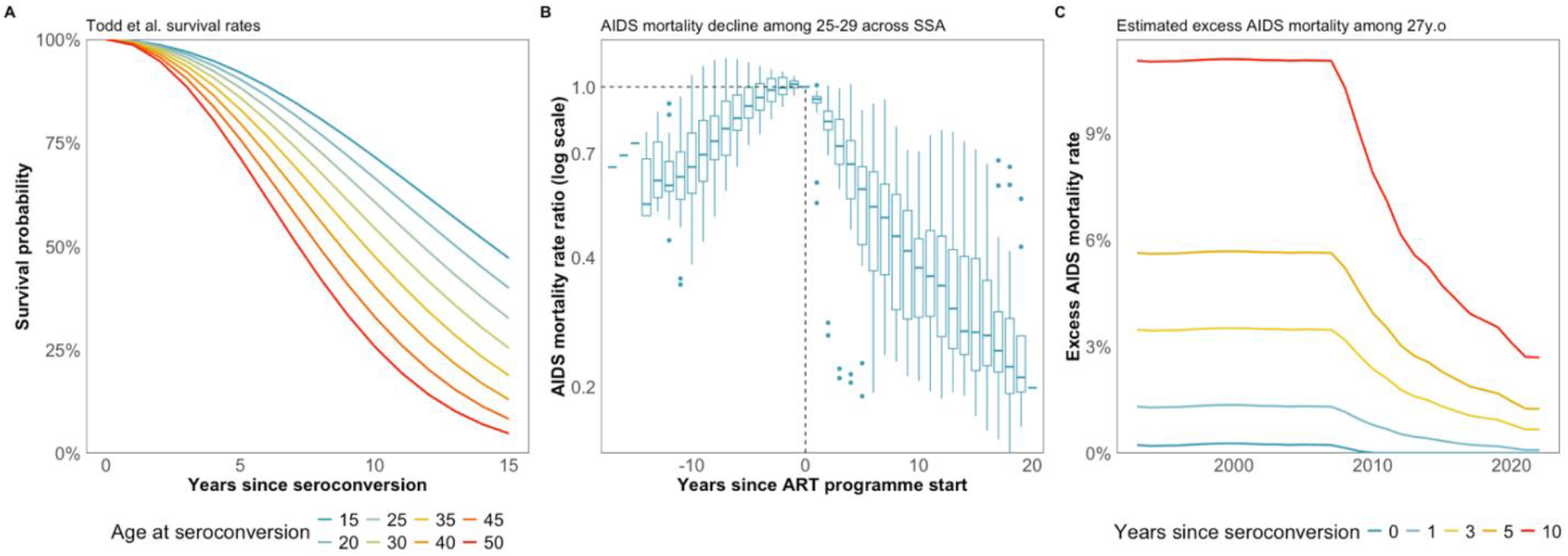
Estimating mortality rates among female sex workers in the ART era – an example for 27 year olds. (A) Mortality rates by age and year since seroconversion in the pre-treatment era from analysis by Todd et al. (B) Decline in AIDS mortality among 25-29 year olds in all Spectrum files in sub-Saharan Africa relative to the year since ART became available in each country (C) Estimated mortality rates since seroconversion among 27 year old sex workers.

### HIV incidence sensitivity analysis

Compared to empirical HIV incidence observations among sex workers aged 18+ (not used in model calibration), We estimated a much higher HIV incidence in 2000 (model: 16.7/100py [6.9-19.1] vs study 8.3/100py [3.4-13.2]) and in 2010 (model: 6.2/100py [2.7-7.2] vs study 1.4/100py [0.3-2.5]). In sensitivity analysis, altering the odds ratio that controls HIV prevalence at sex work initiation (*ι*_*t*_) had a large impact on estimated incidence level and trend (Figure 6.3). Fixing *ι*_*t*_ at 1, 3, 5, or 10 over time all produce similar results that are a poor fit to HIV prevalence data and estimate fluctuating incidence trends that reach very high levels of incidence (30/100py). Empirical incidence may be reconciled with cross-sectional HIV prevalence data by removing the upper bound on *ι*_*t*_ and recruiting women with 30-50% prevalence. Removing the under-enumeration observation model had little effect on HIV incidence trajectories before 2015 whereupon they began to diverge. In 2020, removing the duration observation model decreased HIV incidence by 27% in 2020 (1.4/100py vs 1.9/100py). Simulating an increase in population size to 2% of 15-49 women in 2020 increased incidence by 83% in 2020 (3.4/100py vs 1.9/100py).

### Age and duration distributions

The mean duration of sex work was constant between 3 and 3.5 years from 1993-2008, rose to 5 years by 2015, and then fell to 2.5 years by 2022 (Figure 6.3A). Survey under-enumeration of sex workers at short sex work durations was detected in all surveys since 2005. Sex workers in their first year of selling sex were estimated to be under-enumerated by median 60% (IQR 24-71%) and in their second year of selling sex by 19% (IQR 2-39%; Figure 6.3B). Adjusted for under-enumeration, mean duration decreased by 0.5 years (IQR 0.2-1). The mean age of sex workers has remained unchanged between 1995-2022 at 29.5 ± 1 years (Figure 6.4). The mean age at sex work initiation declined from 28 years old in 1993 to 26 in 2012, and returned to 28 years old by 2019.

**Figure 6_3a:**
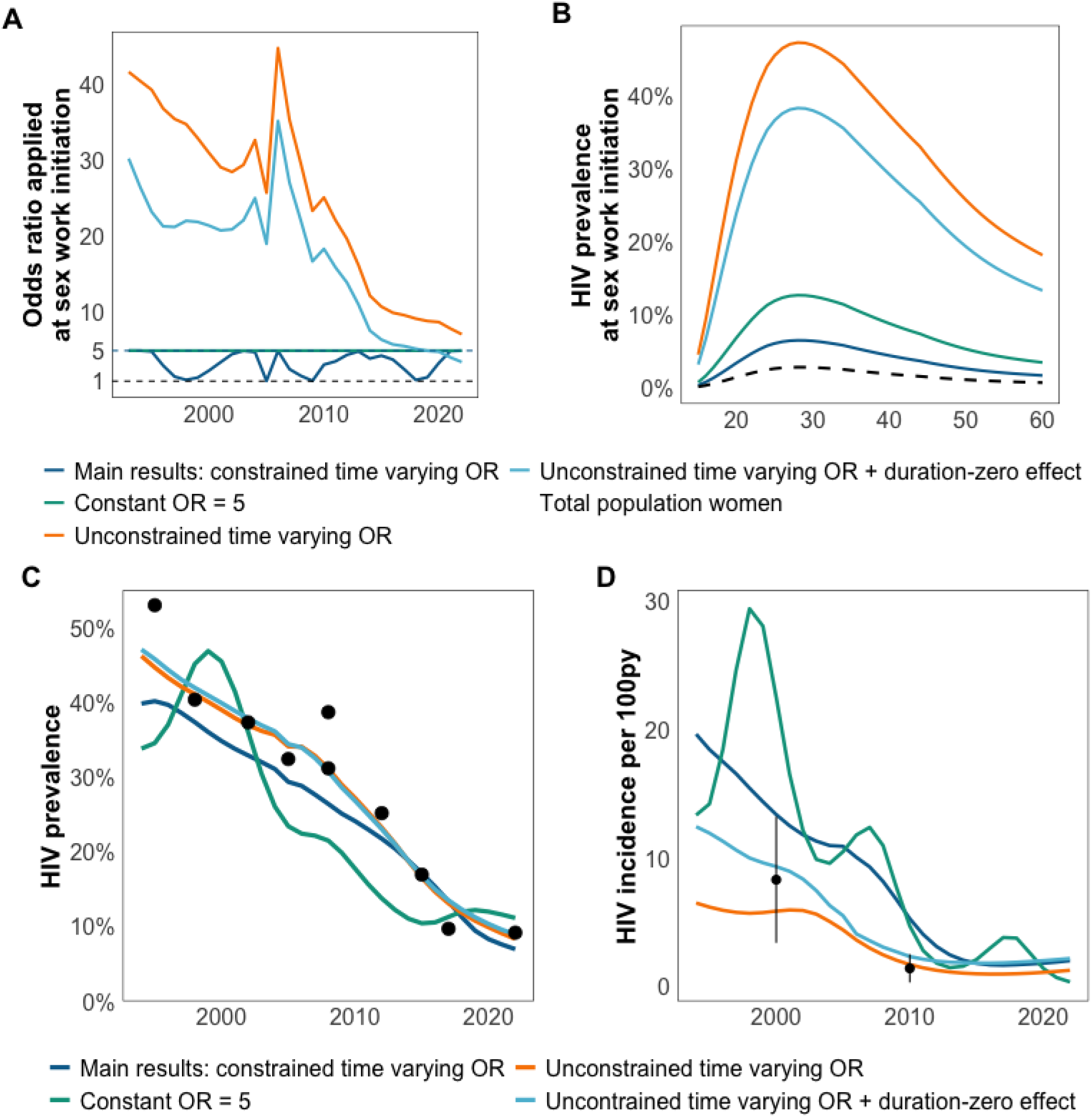
HIV incidence sensitivity analysis. (A) Estimated odds ratio (OR) applied to HIV burden at sex work initiation. OR in all sensitivity models has a minimum bound at OR=1 (black dashed line) and main results (blue) have an upper bound at OR=5. (B) HIV prevalence at sex work initiation in 2000 from each model, compared to HIV prevalence among total population women (black dashed line). (C) Estimated HIV prevalence among ages 15-49 compared to survey data (black points). (D) Estimated HIV incidence compared to empirical observations of HIV incidence (not included in model calibration). Results for constant OR=1, 3, and 10 were similar to constant OR=5 and are not plotted.

**Figure 6_4:**
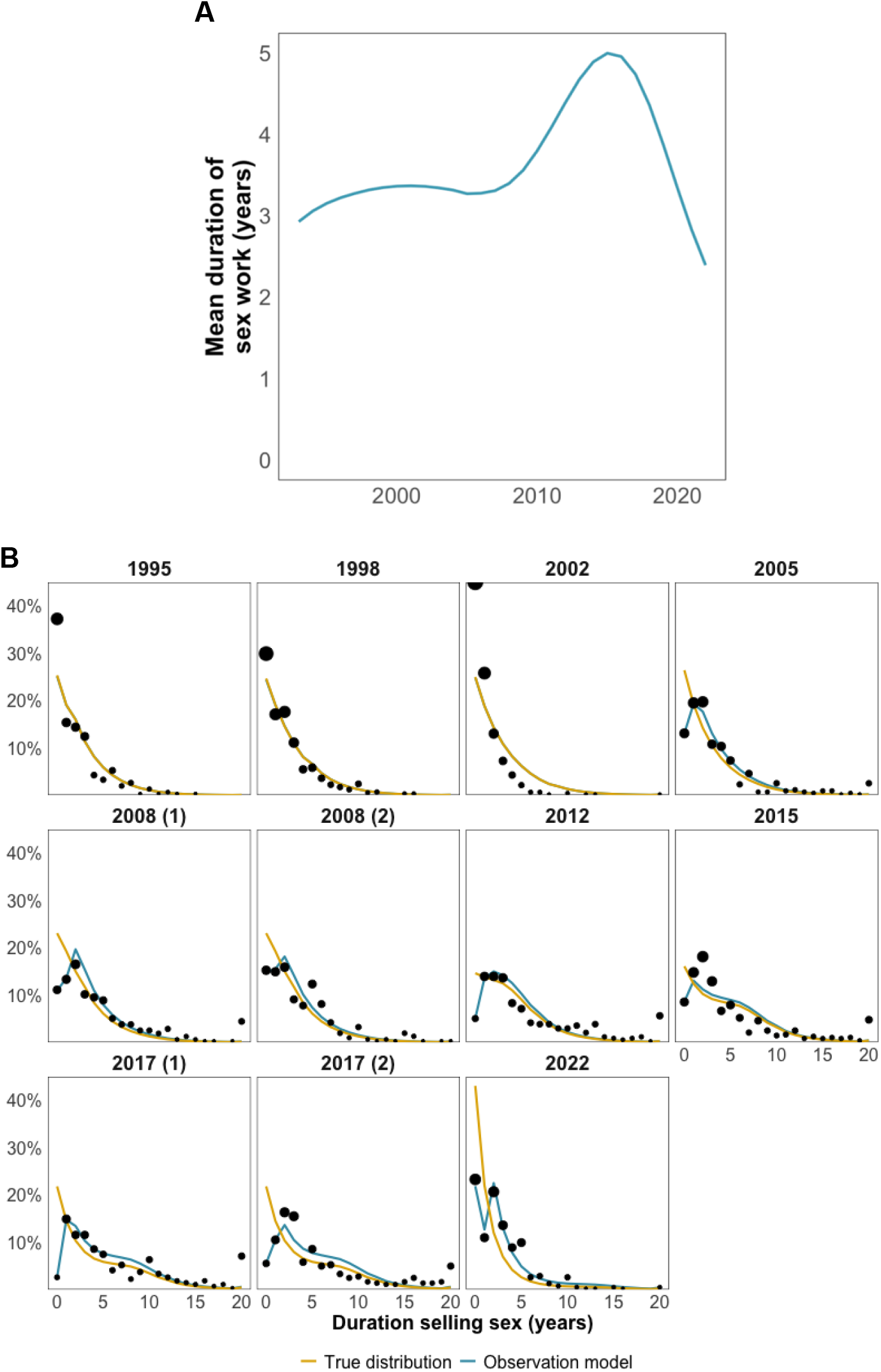
Distribution of duration selling sex. (A) Mean duration of sex work over time. (B) The observation model (blue) includes effects to estimate under-enumeration in the first two years of sex work. Accounting for under-enumeration yields the true distribution (yellow). Estimates shown for survey years.

**Figure 6_5:**
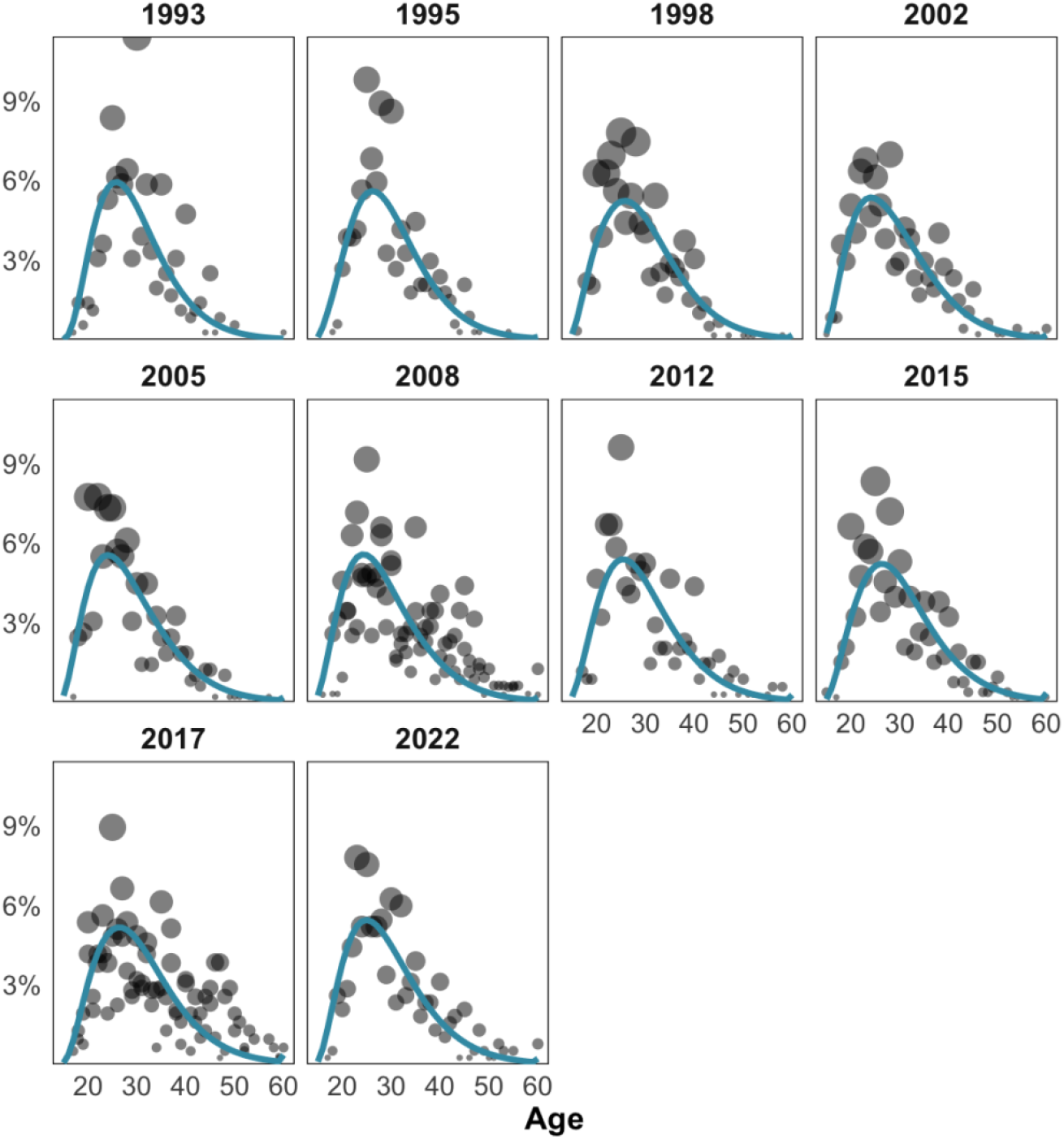
Sex worker age distribution. Estimates (blue) shown in year of survey compared to survey data (black points)

## 1.4 Discussion

We created a statistical model that estimated HIV incidence among female sex workers from serial cross-sectional survey data. It does so without data on CFSW or behavioural parameters of sex work required by full transmission dynamic models. This model could be applied in any setting with one or more surveys among female sex workers. In the Beninese setting, we estimated HIV incidence has declined ten-fold since 1993 to around 2/100py in 2013, whereupon it has plateaued through 2022 (2.6/100py). Incidence in 2022 was 37-fold higher than HIV incidence among total population women aged 15-39 in Western and Central Africa, compared to 20-fold higher as estimated in a recent meta-analysis of empirical observations of HIV incidence among sex workers.^1^

The model fits well to a diverse range of data, including HIV prevalence by age and duration, age and age at sex work initiation, and duration of sex work. Despite this, estimated HIV incidence was much higher than empirical incidence observations not included in model calibration: 2fold higher in 2000 and 4.5fold higher in 2010.^1,19,20^ This is principally driven by survey data that indicate high HIV prevalence among those selling sex for less than 1 year, and little HIV prevalence gradient by duration of sex work. Estimated HIV incidence can be only be aligned with empirical observations if women are recruited at very high HIV prevalence (30-40%) which is is unrealistic in a low prevalence HIV epidemic. Here we will discuss sex work dynamics and biases in surveillance data that may give rise to this difference, and their consequences for interpreting both modelled incidence estimates and empirical incidence observations.

### 1.4.1 Episodic engagement in sex work

Sex work is commonly stratified into ‘commercial’ sex work (selling sex for money only and earning all/most of one’s income from sex work) and ‘transactional’ sex work (selling sex for money, or exchanging for gifts or favours). Transactional sex workers may sell sex episodically, driven by transient conditions (e.g. school holidays, poor harvest), see fewer clients, and experience lower HIV incidence risk. Episodic serial entry and exit from sex work are documented in qualitative accounts, but little quantitative data are available to parameterise this behaviour, and existing model-based approaches have required strong assumptions.^21,22^ This model did not track former sex workers, instead returning sex workers to the total population upon cessation of sex work. If serial engagement and prevalent infection at sex work entry is common, incidence may be lower and less concentrated at short durations than estimated. Model development should look to utilise survey data on client frequency and proportion of income derived from sex work to stratify sex work by non-commercial and commercial sex work, with non-commercial sex workers selling sex for shorter durations with a higher rate of serial cessation and initiation. Granular data on episodic sex work should be viewed as a priority within future surveillance or routine data collection.

### 1.4.2 What is ‘duration’ really measuring?

In this analysis, duration of sex work was directly available from 6 surveys (“for how long have you sold sex?”), and calculated from current age and age at first paid sex in 5 surveys. In the context of episodic sex work, it is unknown whether survey respondents report total duration of sex work or duration of current episode, though it is widely interpreted in secondary analysis and modelling studies as total duration. Of seventeen surveys among female sex workers in SSA that included both a direct question on duration and asked about age at first paid sex, sixteen reported longer durations using age at first paid sex. There is circumstantial evidence from the 2022 survey that total duration may be being misreported in Benin. Benin is a low prevalence epidemic, and we assume that the majority of HIV-positive sex workers acquired HIV within sex work. The 2022 survey conducted viral load testing and found high levels of viral load suppression (VLS) among sex workers selling sex for <1 year. As it would be challenging to initiate sex work, seroconvert, be diagnosed and linked to care, and reach viral load suppression within, on average, six months, it is likely that some of these women had prevalent infections from previous episodes of sex work. Accordingly, their total duration of sex work would be higher than <1 year and HIV prevalence among those truly at <1 year of sex work be lower than that currently observed in survey data.

Average duration from aggregate data is commonly used to parameterise turnover in key population-stratified models, including in Goals-RSM.^23^ HIV incidence estimates are sensitive to sex work duration assumptions: duration determines the rate at which high HIV prevalence FSW are ‘diluted’ by entrants from the low HIV prevalence total population, and in turn, the incidence required to restore observed prevalence. If estimates of total duration are under-estimated, HIV incidence will be over-estimated and as absolute durations of sex work are small, single year changes in average duration could lead to large changes in incidence estimates.

### 1.4.3 Missing women at highest incidence risk

We found evidence for under-enumeration of women selling sex for <1 and 1-2 years from 2005 onwards. Adjusted proportions for women selling sex for <1 year were commonly double survey-observed proportions. Surveys were sampled using hotspot sampling from a peer-generated sampling frame of most-frequented sex work locations, or by clinic-based recruitment. New sex workers may be unaware of these hotspots early in their sex work careers or may have started selling sex informally without attending commercial hotspots. Clinic-based recruitment (including the proposed BBS-Lite^24,25^) and network-based (e.g. RDS) may be further susceptible to under-enumerating individuals at short durations. We estimated that incidence risk declines steeply with duration, similar to incidence estimates among Kenyan sex workers^26^, and programmes may be missing individuals at highest risk soon after, or even before^18^, sex work initiation. Empirical studies may underestimate incidence by under-recruiting women at <1-2 years of sex work where We estimate half of cumulative incidence occurs.

### 1.4.4 Sensitivity to population size assumptions

Sensitivity analysis that doubled population size estimate proportion during the COVID-19 pandemic increased 2020 HIV incidence by 83%. There remains a lack of accurate longitudinal estimates of population size which hinders the production of credible count scale estimates, required for programme planning and commodity purchasing, and impacts on our ability accurately to estimate incidence. This analysis assumed a constant population size proportion. The 2022 survey – conducted shortly after the COVID-19 pandemic - reports a shorter average duration than those preceding it. If the population size proportion is held at 1%, the model obtains a short duration distribution by increasing turnover with a modest increase in incidence to replace the lost prevalence. However, if there is a concomitant increase in population size, the short observed duration is satisfied by an inflow of new entrants with low HIV prevalence, and a large incidence spike is required to reach observed prevalence among those who have sold sex for one year.

### 1.4.5 Mortality estimates and ART coverage

As in catalytic and cohort prevalence methods, the use of external mortality estimates remains a weakness in estimating incidence from cross-sectional prevalence. EPP-ASM stratifies PLHIV by CD4 category to reflect the concentration of AIDS mortality at low CD4 counts, and additionally tracks the number of PLHIV on treatment by CD4 count at initiation and duration on treatment. Data on CD4 count distribution among sex workers are absent from nearly all settings in sub-Saharan Africa. While programme counts of sex workers on treatment are becoming more widely available from key population programmes, interpretation of these data is impaired by programmatic double-counting due to high levels of mobility and multiple service providers, and an unknown fraction of sex workers receiving treatment from total population services.

Without CD4 data and reliable treatment data, sex workers in this model were tracked by time since seroconversion such that estimates of survival could be used in the pre-treatment era. In the treatment era, mortality rates were derived from the average AIDS mortality decline among the total population. Estimates of ART coverage and VLS were available in 2008 and 2022 surveys, 38% and 68% respectively, indicating that FSW had benefitted from the national ART programme similarly to total population women. These could have been used to calibrate the model and inform mortality rates, though the 2008 survey used clinic-based recruitment where ART coverage would be expected to be higher than among all sex workers and conceptually the aim was a parsimonious model that could be applied in settings with weaker surveillance data. Despite evidence of strong programme engagement among FSW, it is possible that approximating mortality among FSW from average decline in AIDS mortality has overstated mortality among FSW and thereby overestimated HIV incidence.

## 1.5 Conclusion

HIV incidence among sex workers in Benin has declined ten-fold since 1993 to around 2.5/100py. Reconciling abundant HIV prevalence data by age and duration should be used to create estimates of HIV incidence in all settings with survey data. These should be used alongside empirical observations and transmission dynamic model estimates of HIV incidence to support HIV prevention programming and targeted provision of pre-exposure prophylaxis.

## Data Availability

All data produced in the present study are available upon reasonable request to the authors

## References

1. Jones HS, Anderson RL, Cust H, et al. HIV incidence among women engaging in sex work in sub-Saharan Africa: a systematic review and meta-analysis. Lancet Glob Health. 2024;12(8):e1244–e1260. doi:10.1016/S2214-109X(24)00227-4

2. Hallett TB, Stover J, Mishra V, Ghys PD, Gregson S, Boerma T. Estimates of HIV incidence from household-based prevalence surveys. AIDS. 2010;24(1):147–152. doi:10.1097/QAD.0b013e32833062dc

3. Hallett TB, Zaba B, Todd J, et al. Estimating incidence from prevalence in generalised HIV epidemics: Methods and validation. Ghys P, ed. PLoS Med. 2008;5(4):611–622. doi:10.1371/journal.pmed.0050080

4. Eaton JW, Brown T, Puckett R, et al. The Estimation and Projection Package Age-Sex Model and the r-hybrid model: New tools for estimating HIV incidence trends in sub-Saharan Africa. AIDS. 2019;33(Suppl 3):S235–S244. doi:10.1097/QAD.0000000000002437

5. Fraser H, Stone J, Wisse E, et al. Modelling the impact of HIV and HCV prevention and treatment interventions for people who inject drugs in Dar es Salaam, Tanzania. J Int AIDS Soc. 2021;24(10). doi:10.1002/jia2.25817

6. Stone J, Fraser H, Walker JG, et al. Modelling the impact of HIV and hepatitis C virus prevention and treatment interventions among people who inject drugs in Kenya. AIDS. 2022;36(15):2191–2201. doi:10.1097/QAD.0000000000003382

7. Stone J, Mukandavire C, Boily M, et al. Estimating the contribution of key populations towards HIV transmission in South Africa. J Int AIDS Soc. 2021;24(1):e25650. doi:10.1002/jia2.25650

8. Geidelberg L, Mitchell KM, Alary M, et al. Mathematical Model Impact Analysis of a Real-Life Pre-exposure Prophylaxis and Treatment-As-Prevention Study Among Female Sex Workers in Cotonou, Benin. J Acquir Immune Defic Syndr. 2020;86(2):e28. doi:10.1097/QAI.0000000000002535

9. Hodgins C, Stannah J, Kuchukhidze S, et al. Population sizes, HIV prevalence, and HIV prevention among men who paid for sex in sub-Saharan Africa (2000–2020): A meta-analysis of 87 population-based surveys. PLoS Med. 2022;19(1):e1003861. doi:10.1371/JOURNAL.PMED.1003861

10. Gregson S, Donnelly CA, Parker CG, Anderson RM. Demographic approaches to the estimation of incidence of HIV-1 infection among adults from age-specific prevalence data in stable endemic conditions. AIDS. 1996;10(14):1689–1697. doi:10.1097/00002030-199612000-00014

11. Williams B, Gouws E, Wilkinson D, Karim SA. Estimating HIV incidence rates from age prevalence data in epidemic situations. Stat Med. 2001;20(13):2003–2016. doi:10.1002/SIM.840

12. Glaubius R, Kothegal N, Birhanu S, et al. Disease progression and mortality with untreated HIV infection: evidence synthesis of HIV seroconverter cohorts, antiretroviral treatment clinical cohorts and population-based survey data. J Int AIDS Soc. 2021;24 Suppl 5(Suppl 5). doi:10.1002/JIA2.25784

13. Mahy M, Lewden C, Brinkhof MWG, et al. Derivation of parameters used in Spectrum for eligibility for antiretroviral therapy and survival on antiretroviral therapy. Sex Transm Infect. 2010;86 Suppl 2(Suppl_2). doi:10.1136/STI.2010.044255

14. Johnson LF, Anderegg N, Zaniewski E, et al. Global variations in mortality in adults after initiating antiretroviral treatment: An updated analysis of the International epidemiology Databases to Evaluate AIDS cohort collaboration. AIDS. 2019;33:S283–S294. doi:10.1097/QAD.0000000000002358

15. Fazito E, Cuchi P, Mahy M, Brown T. Analysis of duration of risk behaviour for key populations: a literature review. Sex Transm Infect. BMJ Publishing Group Ltd. 2012;88 Suppl 2(Suppl 2):i24–i32. doi:10.1136/sextrans-2012-050647

16. Brown T, Peerapatanapokin W. Evolving HIV epidemics: the urgent need to refocus on populations with risk. Curr Opin HIV AIDS. 2019;14(5):337. doi:10.1097/COH.0000000000000571

17. Todd J, Glynn JR, Marston M, et al. Time from HIV seroconversion to death: a collaborative analysis of eight studies in six low and middle-income countries before highly active antiretroviral therapy. AIDS. 2007;21(Suppl 6):S55. doi:10.1097/01.AIDS.0000299411.75269.E8

18. Neufeld B, Cholette F, Sandstrom P, et al. HIV acquisition prior to entry into formal sex work: Inference from next-generation viral sequencing. AIDS. 2023;37(6):987–992. doi:10.1097/QAD.0000000000003484

19. Van Damme L, Ramjee G, Alary M, et al. Effectiveness of COL-1492, a nonoxynol-9 vaginal gel, on HIV-1 transmission in female sex workers: A randomised controlled trial. Lancet. 2002;360(9338):971–977. doi:10.1016/S0140-6736(02)11079-8

20. Diabaté S, Chamberland A, Geraldo N, Tremblay C, Alary M. Gonorrhea, Chlamydia and HIV incidence among female sex workers in Cotonou, Benin: A longitudinal study. PLoS One. 2018;13(5):e0197251. doi:10.1371/JOURNAL.PONE.0197251

21. Kerrigan D, Mbwambo J, Likindikoki S, et al. Project Shikamana: Baseline Findings From a Community Empowerment–Based Combination HIV Prevention Trial Among Female Sex Workers in Iringa, Tanzania. J Acquir Immune Defic Syndr. 2016;74(Suppl 1):S60. doi:10.1097/QAI.0000000000001203

22. Jesse Knight A, Wang S, Mishra S, et al. Adjusting for hidden biases in sexual behaviour data: a mechanistic approach. medRxiv. Published online August 20, 2023:2023.08.16.23294164. doi:10.1101/2023.08.16.23294164

23. Stover J, Glaubius R, Teng Y, et al. Modeling the epidemiological impact of the UNAIDS 2025 targets to end AIDS as a public health threat by 2030. Rosen S, ed. PLoS Med. 2021;18(10):e1003831. doi:10.1371/journal.pmed.1003831

24. Gogia M, Ruadze E, Kasrashvili T, et al. Piloting a simplified bio-behavioural survey methodology, the BBS-Lite, among people who inject drugs in Georgia. Int J Drug Policy. 2024;144(Pt 1). doi:10.1016/J.DRUGPO.2024.104326

25. UNAIDS, WHO. The bio-behavioural survey “lite”: a methodology for monitoring programmes providing HIV, viral hepatitis and sexual health services to people from key populations. Implementation tool. Published online 2024. Accessed September 5, 2025. https://www.who.int/about/policies/publishing/copyright.

26. McKinnon LR, Izulla P, Nagelkerke N, et al. Risk Factors for HIV Acquisition in a Prospective Nairobi-Based Female Sex Worker Cohort. AIDS Behav. 2015;19(12):2204–2213. doi:10.1007/S10461-015-1118-7/FIGURES/1

